# Cohort profile: working age adults accessing secondary mental health services in South London (UK) and benefits – a data linkage of electronic mental health records and benefits data

**DOI:** 10.1101/2023.10.18.23297221

**Authors:** Ava Phillips, Ray Leal, Amelia Jewell, Ira Madan, Johnny Downs, Matthew Broadbent, Matthew Hotopf, Sarah Dorrington, Nicola T. Fear, Sharon A.M. Stevelink

## Abstract

This study describes a cohort of working age adults from a novel data linkage. The data linkage is comprised of routine electronic mental health records from The South London and Maudsley (SLaM) NHS Foundation Trust and benefits records from The Department for Work and Pensions (DWP). The data linkage spans 15 years, with SLaM data covering the period 2007-2019 and DWP data covering the period 2005-2020. The linkage was established to investigate the interrelationships between benefit receipt and mental health (including mental health service utilisation) in patients accessing secondary mental health services in South London, UK. Clinical data from SLaM includes information on sociodemographics, diagnoses, assessments, treatments, interventions, and service interaction. Benefits data from DWP provides information on benefit types received, spell dates and entitlement amounts, work capability assessments and work programme access.

This cohort included all working age adults (aged 18-65 years) referred to/receiving treatment from SLaM services between January 2007 and June 2019. We report the sociodemographics and psychiatric diagnoses of n=150,348 working age adults, as well as the types of benefits received. We showed that 78.3% of working age adults received a benefit relating to unemployment or work-related income, sickness or disability, or a housing benefit, and that 68.0% of this group had a recorded primary psychiatric diagnosis. The most commonly received diagnoses were alcohol related disorders and substance related disorders (17.2%), followed by mood (affective) disorders (16.5%). We also observed that nearly 70% of claimants who had received more than one benefit were residing in the two most deprived areas across the South London boroughs covered by SLaM.

## Introduction

In the UK, mental disorders are one of the most frequently reported working age disabilities and/or health conditions and reasons for claiming government unemployment, work-related income, sickness, and disability related benefits 1]. It was reported in a health and disability green paper by the Department for Work and Pensions (DWP) that spending on benefits for people with disabilities and people with long-term health conditions is at its highest, with over £30 billion being spent in 2019/20 on working age health and disability benefits 2]. This is predicted to increase to £31 - 40 billion by 2025/26. In the UK, the Department for Work and Pensions (DWP) provides social security for those in need, known as “benefits”. Different types of benefits are provided depending on people’s needs and circumstances e.g., unemployment benefits for people who are out of work, benefits for those on a low income, or sickness and disability benefits for people with a disability, long-term illness, or mental disorder 3]. The standard entitlement amounts for each benefit differ and are also dependent on people’s circumstances such as their income, whom they live with, if they are married and how many children they have 4]. There have been substantial changes to the UK benefits system in the past two decades, specifically the introduction of “Universal Credit” (UC) which is a benefit provided for people who are out of work as well as for those who are in work but on a low income and require extra support. It replaced 6 other benefits (Housing Benefit, Income Support, income-based Jobseeker’s Allowance, income-related Employment and Support Allowance, Child Tax Credit and Working Tax Credit). Although these are being phased out, some people still receive these benefits, known as “legacy benefits”, as they have not yet been transferred onto UC. UC was introduced as part of the *Welfare Reform Act 2012*, in order to reduce the complexity of claiming various different benefits 5]. Another key reform is the change over from Disability Living Allowance (DLA), a benefit for people who need support with mobility or care costs, to Personal Independence Payment (PIP). PIP is a non-means tested benefit, meaning a person’s income is not relevant to entitlement of receiving this benefit, and is similarly provided to help with extra costs due to having ill health or a disability, whether the claimant is in or out of work. Individuals are required to undergo a points-based assessment to determine how their condition impacts their functional ability 6].

Despite the notion that these reforms support the government’s drive to help people feel motivated to obtain and remain in employment 5], research indicates evidence of concern for those with mental disorders in particular as a result of these changes. Research has shown that people experience further distress and, therefore, a reduced likelihood of employment 6
7].

No UK-wide dataset currently provides combined individual level, routinely collected mental health service data and benefits data. It is vital that research into the provision and ongoing needs of people with a disability, long-term illness or mental disorder is carried out to inform policy on how best to support some of the most vulnerable members of society. This paper aimed to establish and describe a working age cohort within a record-level linkage which provides an opportunity to better understand benefit receipt among those with mental disorders, the patterns of engagement with both National Health Service (NHS) secondary mental health services and the benefits system, and the impact of changes to the benefits system over the past two decades. We present sociodemographic characteristics and data on psychiatric diagnosis and benefit receipt among working age adults accessing secondary mental health care services and benefits in Southeast London.

### Ethics approval

The South London and Maudsley (SLaM) NHS Foundation Trust and Department for Work and Pensions (DWP) data linkage resulted in a linked data set of longitudinal data from SLaM electronic mental health records and DWP benefits data. All data used is de-identified. The use of SLaM mental health records data for research purposes received full ethical approval from the NHS Research Ethics Committee (Oxford South Central ref 18/SC/0372). Ethical approval for the data linkage was granted from the Oxford South Central REC (Ref: 22/SC/0400), and the data linkage was legally permitted by the Health Research Authority Confidentiality Group (CAG) under Section 251 of the NHS Act 2006 (ref 17/CAG/0055).

The Maudsley Biomedical Research Centre (BRC) data linkage service user and carer advisory group is a Patient and Public Involvement and Engagement (PPIE) group containing members with lived experience of mental disorders or caring for someone with a mental disorder who has accessed mental health services. Members were involved in early discussions around the data linkage and were supportive of the linkage. In addition, two PPI members have joined our research team and are regularly involved in discussions regarding projects using the linked data. Consultation and discussion with all PPI members will continue as work progresses 8].

### Cohort description

Mental health data was extracted from the electronic health care records of the SLaM NHS Foundation Trust; one of Europe’s largest providers of secondary mental health services, providing services to the South London boroughs of Lambeth, Lewisham, Southwark, and Croydon, covering a catchment area of over 1.3 million residents 9]. SLaM provides specialist secondary mental health services for a number of disorders, including eating disorders and psychosis related disorders, as well as outpatient services for low level common mental disorders (for example, Improving Access to Psychological Therapy (IAPT) services). Patients who accessed SLaM services vary in their level and method of contact with services, for example patients may a have one-off presentation or referral from a general practitioner (GP), or ongoing hospitalisations and treatment. Patients included in the current cohort are those who had accessed either SLaM secondary mental health services (some of whom will have also accessed IAPT services). Those who only accessed IAPT services were excluded from this cohort, as the focus was on patients who were likely to meet clinical thresholds for a mental disorder. The Maudsley Biomedical Research Centre (BRC) Clinical Records Interactive Search (CRIS) system was established to provide a service-user led governance framework, that could safely and securely de-identify clinical data from SLaM’s electronic health records for research purposes 10].

Benefits data comes from the DWP. DWP are responsible for policy implementation relating to most benefits in the United Kingdom. The linkage of the two datasets took place in 2020 using an ad hoc deterministic approach, achieving a linkage rate of 92.3% [see Reference 11 for full details].

This profile paper reports on a cohort of patients of working age who accessed SLaM services between 1^st^ January 2007 and 30^th^ June 2019. We defined working age as those between the ages of 18-65 years, including those aged 18 years and above at SLaM window start date (January 2007) and those aged 65 years and below at SLaM window end date (June 2019) (Figure 1). This is because people in the UK turning age 66 years were eligible for State Pension and this was generally considered as the standard retirement age 12].

We report data on working age patients who had accessed SLaM (n=150,348) and some of them had received a benefit relating specifically to unemployment or work-related income, sickness or disability, or Housing Benefit, which are referred to as benefits of interest.

### Cohort timeline

The linkage provides data covering a 15-year window, with benefits data starting from 1^st^ January 2005 until 30^th^ June 2020, and data from SLaM covering the period 1^st^ January 2007 until 30^th^ June 2019. Due to the longitudinal and administrative nature of the linkage, data can exist for people, in relation to both mental health data and benefits data at multiple timepoints. For some, data on mental health and benefits exist together throughout the period under study. For others, there may be only one instance for both mental health and benefits data. It should be noted that the CRIS data is ‘live’, so for each new project specific data extraction using the linked data, details can vary as the SLaM patient journey is ongoingly edited by health professionals.

### Cohort measures

#### Mental health electronic record data

All patient level information within the CRIS system was pseudonymised, including structured fields and unstructured text from clinical notes. Data included individual level socio-demographic characteristics (e.g., sex, month and year of birth, date of death, ethnicity, marital status, neighbourhood deprivation) and time variant clinical data such as psychiatric diagnosis, assessments, mental health treatment (medication and therapies), and engagement with SLaM services including admissions, recorded attendance, and adherence to treatments. Note that it is possible patients received different psychiatric diagnoses over time; the psychiatric diagnosis reported in the tables refers to the first instance of a psychiatric diagnosis received within the SLaM data window (January 2007-June 2019).

##### Benefits data

Socio-demographic data included sex, date of death and truncated postcode. Time variant data included dates of benefit receipt. The benefits data were derived from various benefits sources, for example, the “National Benefits Database” (NBD) contains certain benefit types, many of which were legacy benefits, whilst “Universal Credit” (UC) and “Personal Independence Payment” (PIP) was extracted from different administrative data sources. There were also separate benefit “flags” for Pension Credit and Housing Benefit (HB) within the NBD database. NBD benefits include Incapacity Benefit (IB), Carer’s Allowance, Income Support (IS), Jobseeker’s Allowance (JSA), Attendance Allowance, Retirement/State Pension, Disability Living Allowance (DLA), Severe Disablement Benefit (SDA), Widow’s Benefit, Passported Incapacity Benefit, Bereavement Benefit and Employment and Support Allowance (ESA). Data contained benefit spell dates and payment entitlements. A benefit spell indicated a period of time a person was receiving a benefit without a break. There was also information on work-related interventions provided by DWP, as well as work capability assessments (WCAs), which are used to determine someone’s capacity for work and entitlement to ESA. The WCA was introduced as part of a wider benefit reform in 2008 13].

For the purpose of this paper, we only examined benefits relating to unemployment or work-related income, sickness or disability, or HB, all of which may be relevant for working age adults who have accessed secondary care services for their mental health. These benefits of interest include UC, PIP, HB, IB, IS, JSA, DLA, SDA, and ESA. Although not an unemployment, work-related income, sickness or disability related benefit, HB was included in the benefits of interest, as this is a key benefit that is only received alongside another benefit and is crucial in providing a fuller picture of someone’s benefits situation.

Table 1. presents key variables. This is not an exhaustive list of every specific benefit related variable in the benefits data, or all clinical variables held within the CRIS data.

**Table 1.**
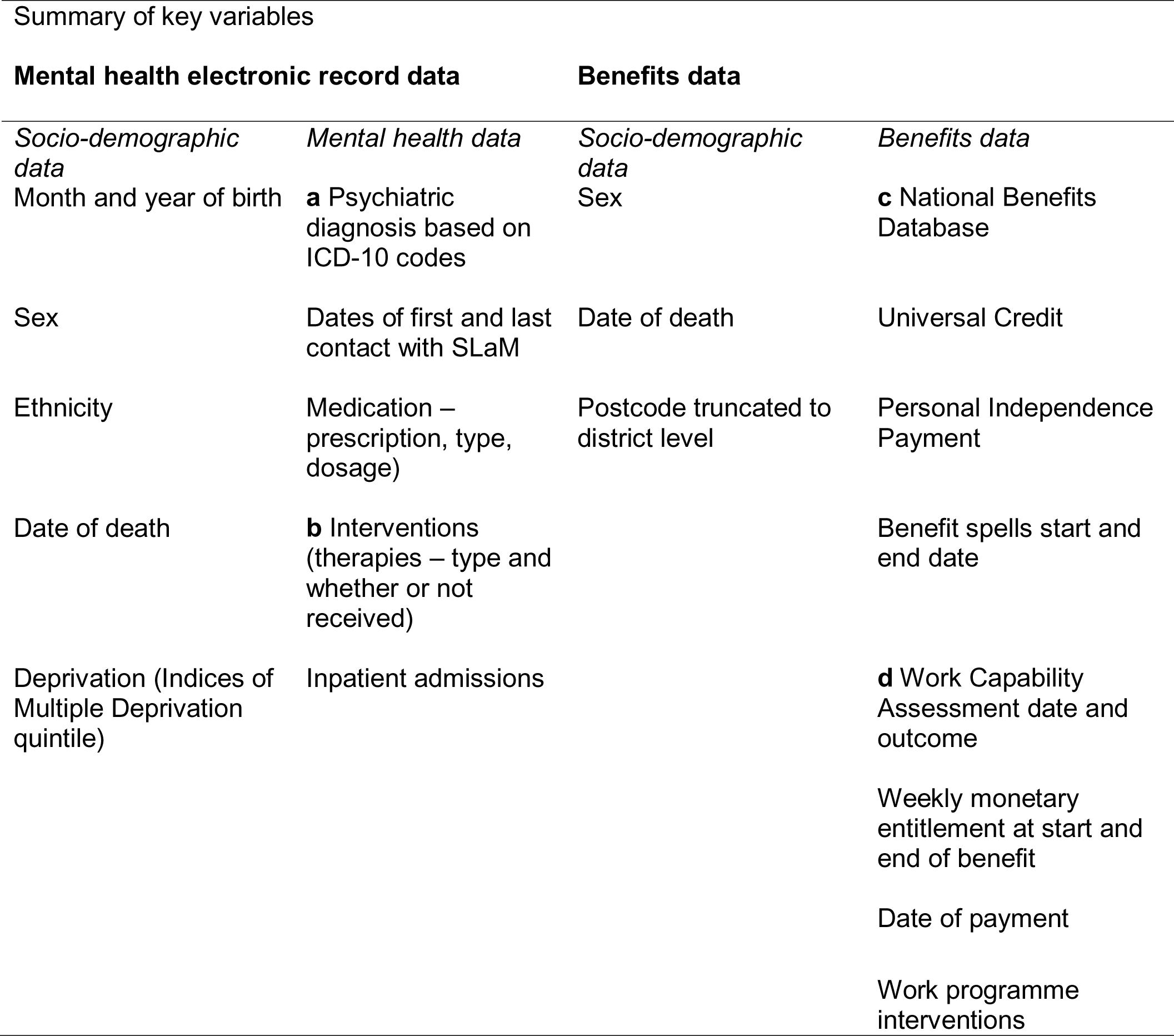
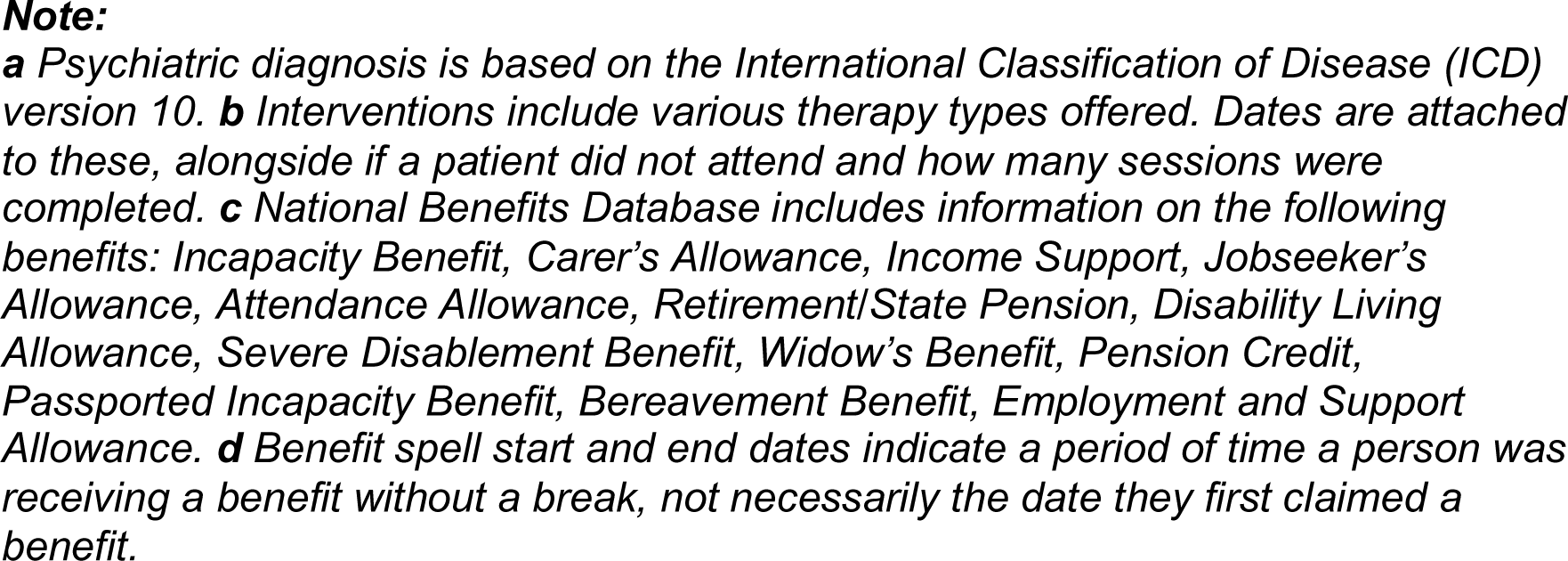
Key variables from SLaM mental health electronic record data and benefits data from DWP.

This data enables us to look at the on and off flows of benefit receipt and mental health utilisation among all patients. We can determine how long people are receiving a benefit(s) for, entitlement amounts, as well assessment dates and outcomes. Data on interventions from DWP provide information on a variety of different intervention types that have been offered to patients, including but not limited to “Skills for Work”, “Job Search Support Centre” and “Job Club”, as well as the start and end dates of when these opportunities have been provided and the “end reason” such as “Found Work”.

## Findings to date

We have reported initial findings on the linkage process between SLaM and DWP records and described the linkage process and sensitivity analysis exploring factors associated with linkage success between SLaM patients and their DWP records. 10]. Of the characteristics studied, female sex, ethnic minority status, being aged between 21-60 years relative to other ages below or above, and not having a recorded primary psychiatric diagnosis were associated with non-successful linkage 11].

For this cohort profile report, we explored the socio-demographic and clinical characteristics of working age adults who were either receiving or not receiving benefits of interest. Of the 150,348 working age adults who were successfully linked, 78.3% received a benefit of interest (n=117,683). Of this group, 68.0% had a recorded primary psychiatric diagnosis. Of those who either did not receive a benefit at all or received a benefit that was not of interest (e.g., State Pension) (n=32,665), 58.1% had a recorded primary psychiatric diagnosis (Table 2).

**Table 2:**
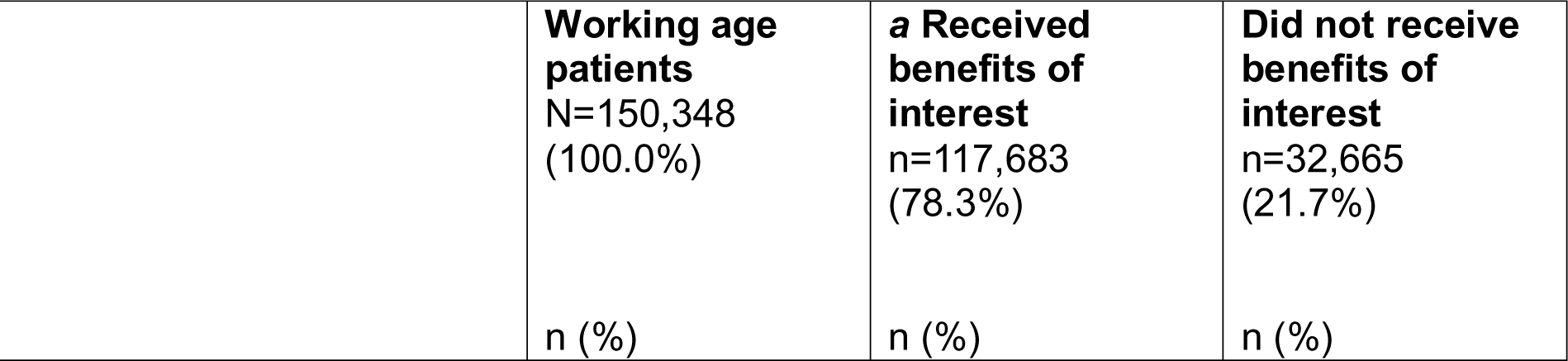
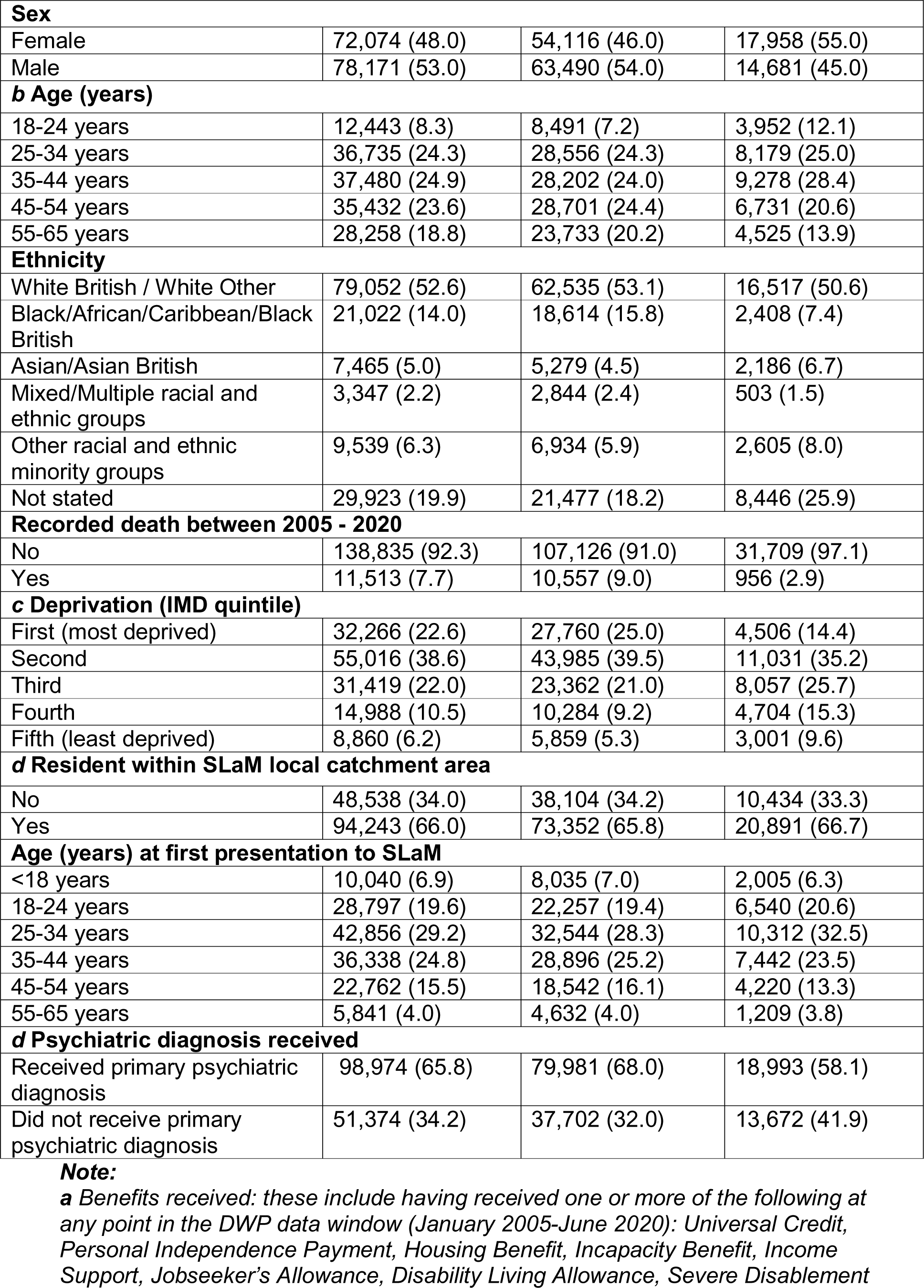
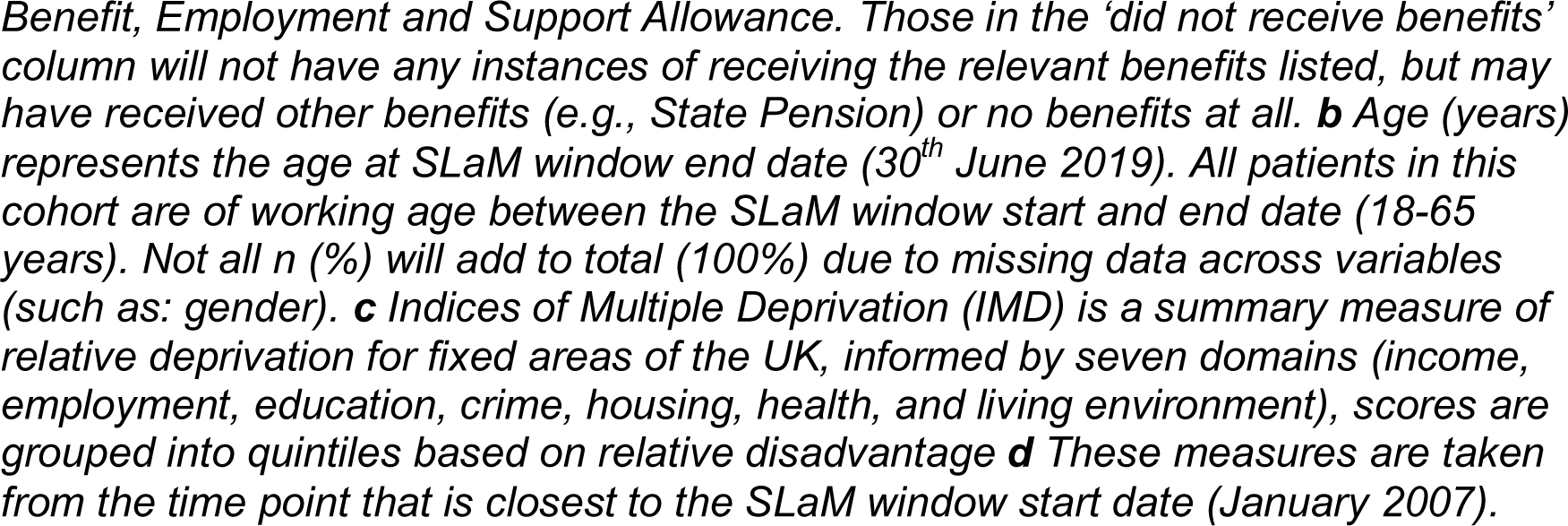
Socio-demographic characteristics of working age patients who have received or not received a benefit related to unemployment, sickness, disability, housing, or income support.

It should be noted that the group of patients who were of working age but did not receive a benefit of interest (n=32,665) included: those who had engaged with DWP as evidenced by an entry in benefits records but did not receive any benefits (n=273), those who had no entry in benefit records (n=31,538), and those who engaged with DWP and received benefits that were not of interest (e.g. State Pension) (n=854). Sensitivity analyses were conducted to assess any key differences between these groups and showed no notable differences.

Table 3 shows that a much higher percentage of patients aged over 24 received more than one benefit compared to those between 18-24 years. This result is likely a consequence of patients in older age brackets having a higher propensity to seek housing benefits. Moreover, belonging to an older age group may suggest an extended duration of benefit receipt, indicating a potential combination of receiving legacy and newer benefits, alongside an elevated need that contributes to a higher frequency of benefit claims.

**Table 3:**
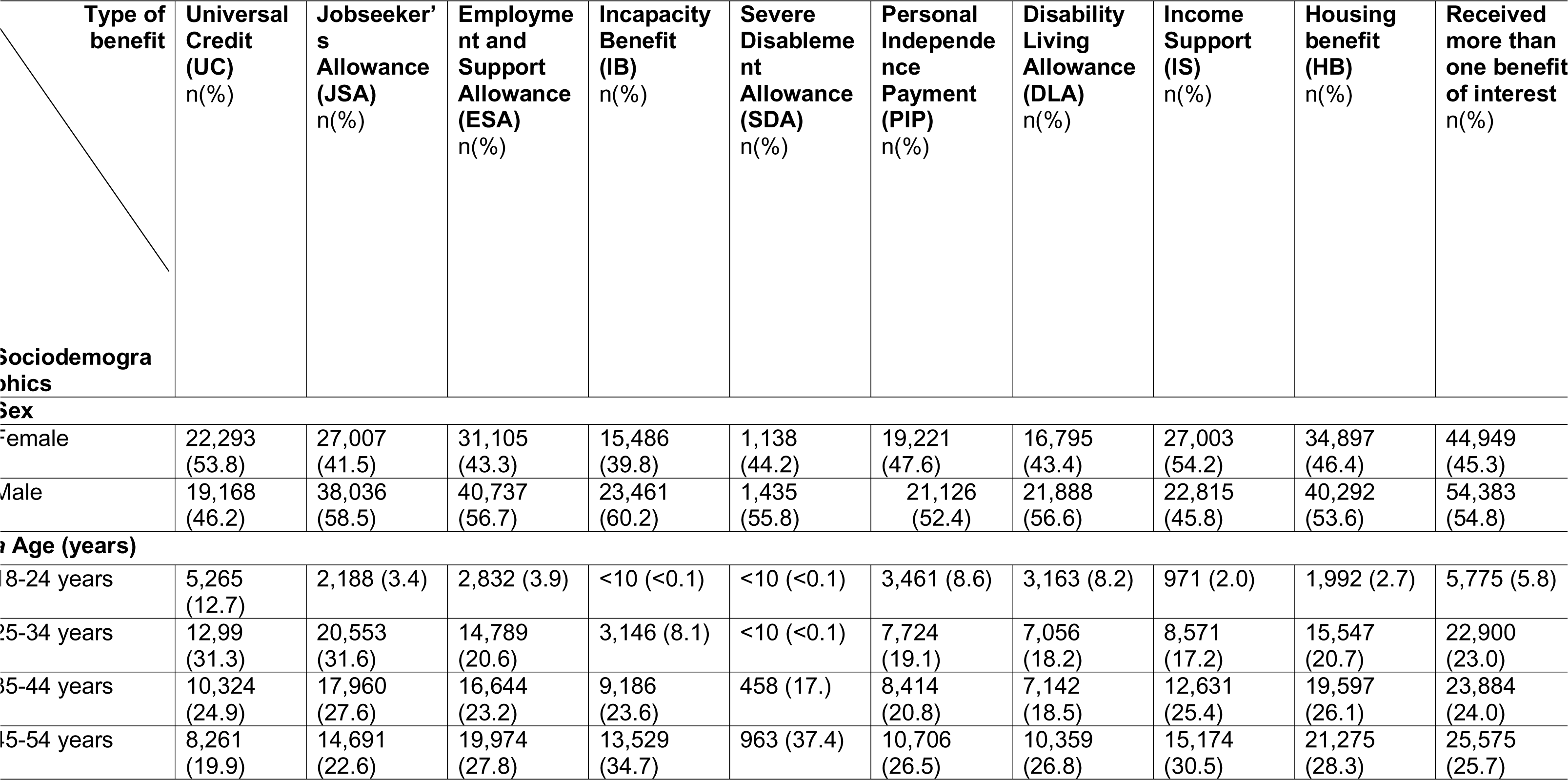
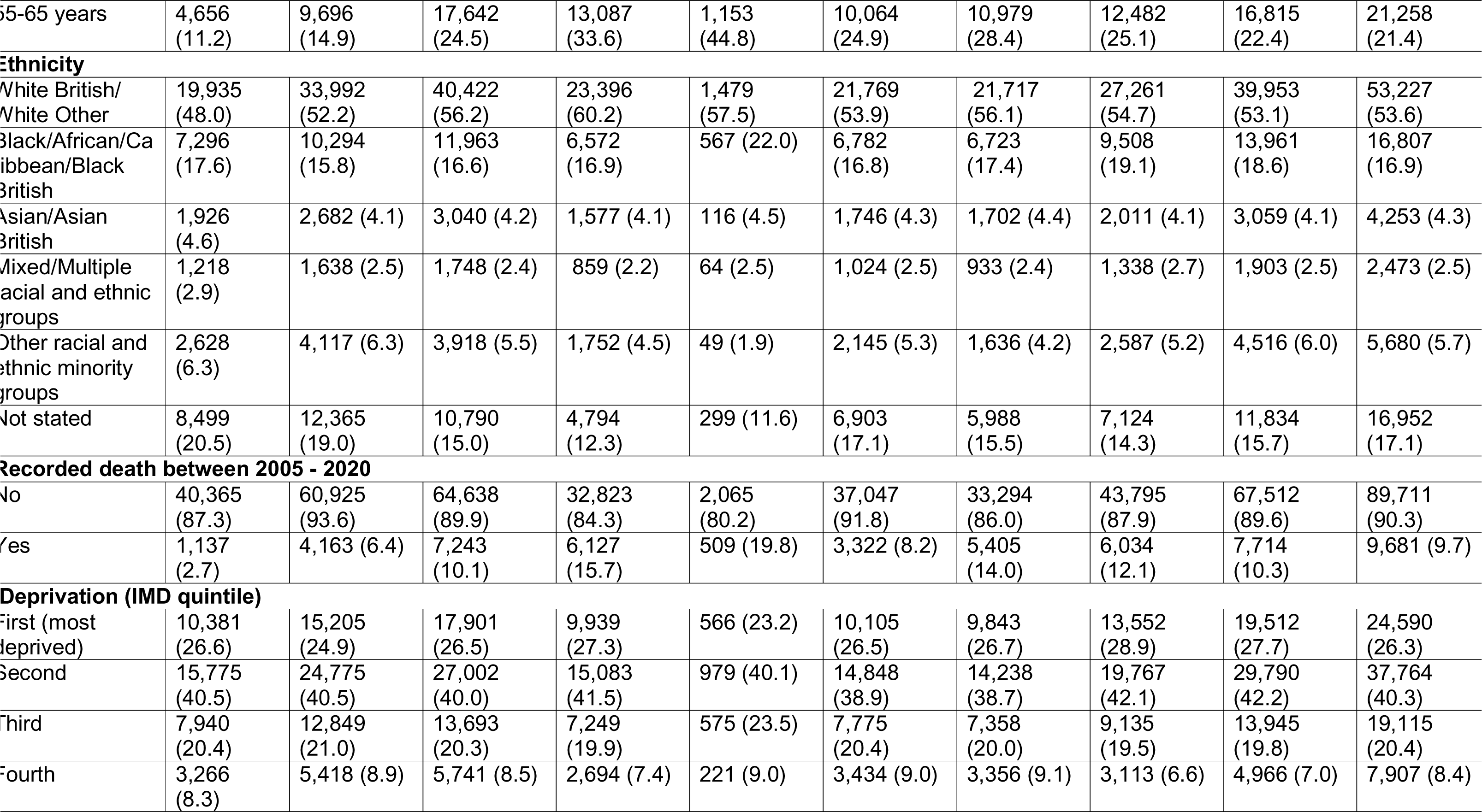
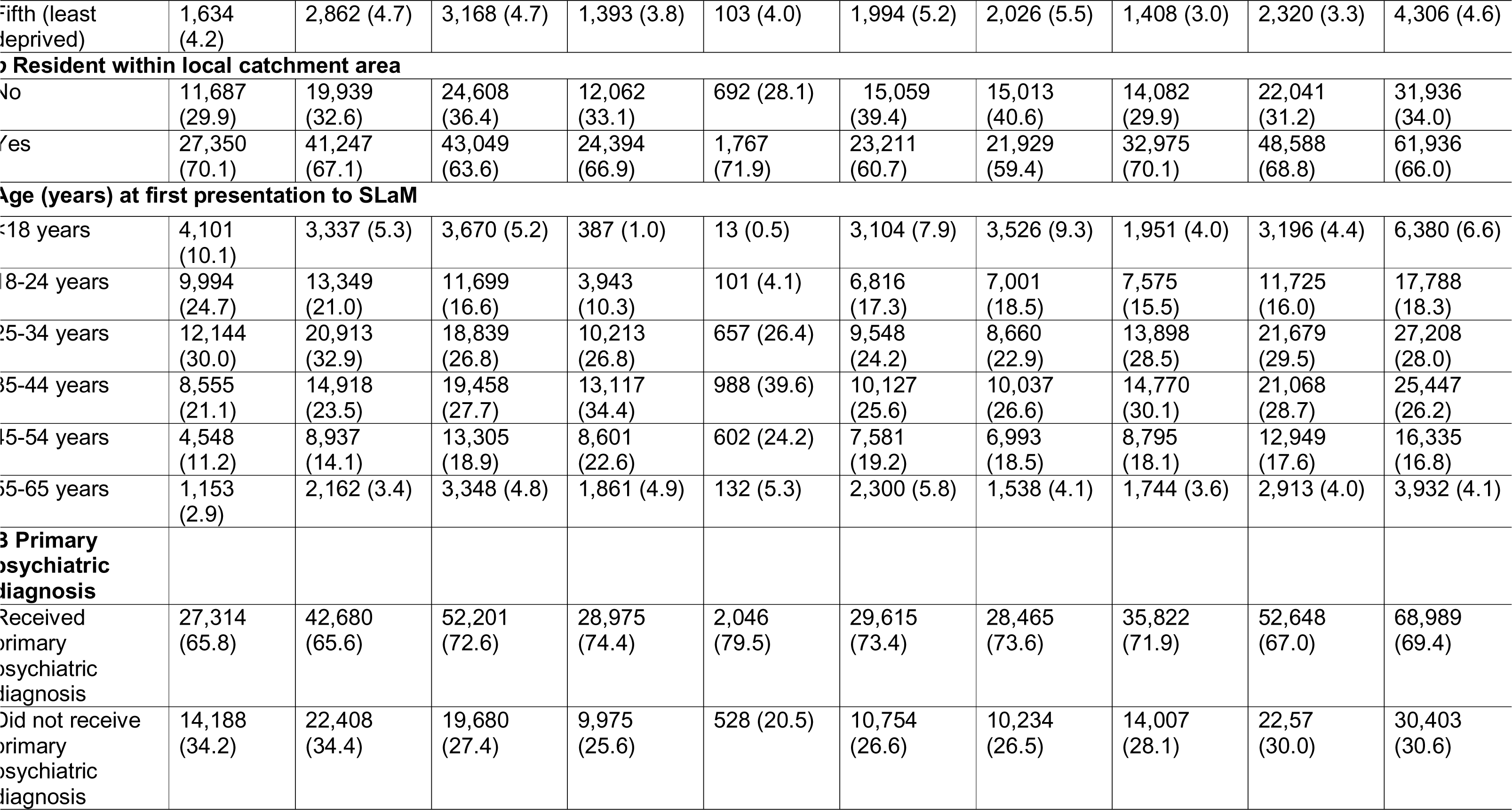
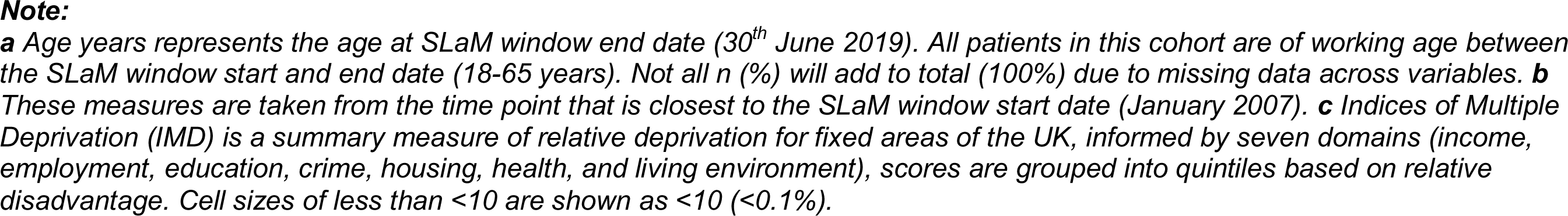
Socio-demographics of working age patients receiving benefits related to unemployment, sickness, disability, housing or income support.

A higher percentage of those within categories “White British” and “White Other” received more than one benefit (53.6%), relative to other ethnic groups. The distribution of ethnic groups corresponds to the demographic distribution in the relevant boroughs. People living in the first and second most deprived areas made up nearly 70% of claimants that had received more than one benefit. This indicates patients residing in more disadvantaged areas need additional support, highlighting the importance of further investigation into this particular group.

A much higher percentage of those with a primary psychiatric diagnosis received more than one benefit (69.4%) compared to those who had not received a primary psychiatric diagnosis (30.6%). This finding aligns with earlier reports, indicating that a prevalent working-age disability and common reason for claiming unemployment and sickness-related benefits is a mental health disorder. Consequently, individuals with a formal psychiatric diagnosis within this group may be more likely to need and receive increased benefits.

Of patients receiving a benefit of interest with a recorded primary psychiatric diagnosis, the most common diagnoses were alcohol related disorders (F10) and substance related disorders (F11-F19) (17.2%), followed by mood (affective) disorders (F30-F39) (16.5%), in particular a single episode of major depressive disorder (F32), and those with anxiety, dissociative, stress-related, somatoform and other nonpsychotic mental disorders (F40-F48) (12.6%) (Table 4).

**Table 4:** Diagnostic categories of working age patients who have received or not received a benefit related to unemployment, sickness, disability, housing, or income support.

| Benefit receipt | Received benefits of interest<br>n=117,683 |
| --- | --- |
|  | n (%) |
| **Recorded first primary psychiatric diagnoses (ICD-10 codes and description)** |  |
| **Schizophrenia, schizotypal, delusional disorders, and other non-mood psychotic disorders (F20-F29)** |  |
| *Total n = 10,586/117,683 (9%)* |  |
| Schizophrenia (F20) | 5,752 (54.3) |
| All other psychosis (e.g., unspecified nonorganic psychosis) (F21-F29) | 4,834 (45.7) |
| **Mood (affective) disorders (F30-F39)** |  |
| *Total n = 19,401/117,683 (16.5%)* |  |
| Major depressive disorder, single episode (F32) | 12,058 (62.2) |
| All other mood affective disorders including bipolar (F30, F31, F33, F34, F38, F39) | 7,343 (37.8) |
| **Anxiety, dissociative, stress-related, somatoform and other nonpsychotic mental disorders (F40-F48)**<br>*Total n = 14,838/117,683 (12.6%)* |  |
| Reaction to severe stress, and adjustment disorders (F43) | 6,110 (41.2) |
| Phobic anxiety disorders (F40), obsessive compulsive disorders (F42), dissociative and conversion disorders (F44), somatoform disorders (F45), Other non-psychotic mental disorders (F48) | 4,808 (32.4) |
| Other anxiety disorders (F41) (e.g., panic disorders, generalised anxiety disorders, mixed anxiety, and depressive disorders) | 3,920 (26.4) |
| **Mental and behavioural disorders due to psychoactive substance use (F10-F19)**<br>*Total n = 20,095 /117,683 (17.1%)* |  |
| Alcohol related disorders (F10) | 8,991 (44.4) |
| Substance related disorders (F11-F19) | 11,104 (55.6) |
| **Behavioural syndromes associated with physiological disturbances and physical factors F50-F59**<br>*Total n = 2,482 /117,683 (2.1%)* |  |
| Eating disorders (F50) and all other behavioural syndromes (F51, F53, F54, F55, F59) | 2,482 (100.0) |
| **Disorders of adult personality and behaviour (F60-F63)**<br>*Total n = 2,856 /117,683 (2.4%)* |  |
| Personality disorders and adult behavioural disorders (F60-F63) | 2,856 (100.0) |
| **Other disorders - intellectual disabilities (F70-F79), pervasive and specific developmental disorders (F80-F89), and behavioural and emotional disorders with onset usually occurring in childhood and adolescence (F90-F98)**<br>*Total n = 9723/117,683 (8.3%)* |  |
| Intellectual disabilities (F70-F79) | 1,851 (19.0) |
| Autism and Asperger's related development disorders (F84) | 2,385 (24.5) |
| ADHD related disorders (F90) | 4,089 (42.1) |
| Other disorders (F80, F81, F83, F88, F89, F91, F92, F93, F94, F95, F98) | 1,398 (14.4) |
| **No recorded primary psychiatric diagnosis**<br>*Total n = 37,702 /117,683 (32.0%)* | 37,702 (100.0) |

**Note:**
*Benefits of interest include having received one or more of the following at any point in the DWP data window (January 2005-June 2020): Universal Credit, Personal Independence Payment, Housing Benefit, Incapacity Benefit, Income Support, Jobseeker's Allowance, Disability Living Allowance, Severe Disablement Benefit, Employment and Support Allowance. All patients in this cohort are of working age between the SLaM window start and end date (18-65 years). The following diagnostic categories were excluded as they were not of interest for the purpose of this study and/or had very small numbers: F01 to F09: Mental disorders due to known physiological conditions, F52 Sexual dysfunction not due to a substance or known physiological condition, F64 Gender identity disorders, F65 Paraphilias, F66 Other sexual disorders, F68 Other disorders of adult personality and behaviour, F69 Unspecified disorder of adult personality and behaviour. If a patient received a first psychiatric diagnosis of FXX or F99, the next F\* code diagnosis they received closest to window start date of the SLaM data window is considered their first psychiatric diagnosis.*

Future research using the linked data will explore both the level of interaction with secondary mental health services and the duration of benefits received over time across different groups (e.g., across diagnostic groups and by socio-demographic factors), as well as assessing temporality between receiving a mental health diagnosis or accessing secondary mental health services and receiving a benefit. The data linkage also presents opportunities to look at the impact of the benefit reforms on mental health outcomes (e.g., mental health severity and diagnosis over time).

## Strengths and limitations

This data linkage is the first of its kind to link electronic mental health records with benefits data in the UK, providing in depth data spanning a 15-year window and opportunity to explore the complex longitudinal relationships between benefit receipt among a substantial number of adults accessing secondary mental healthacross areas of Southeast London. The mental health data provides rich clinical information on psychiatric diagnoses, illness severity, assessment, history of appointments and treatments and interventions received. Benefits data provides substantial data such as type, level, dates of and entitlement amounts of benefits received, WCAs and work-related interventions offered.

Previous research reporting on mental health and benefit receipt has been limited in detail, as well as the methods used. The Adult Psychiatric Morbidity Survey (APMS) (2014) 14] reported that mental disorders are more common among those who were unemployed and receiving an out of work benefit, namely ESA, due to poor health and disability. The current data linkage presents extensive data recorded by health service professionals, as opposed to self-report, and provides more detailed benefits related information. Unlike cross-sectional studies, the longitudinal nature of the linkage presents an opportunity to understand ongoing changes and potential challenges in the provision of mental health services and benefits, as well as capture individuals using mental health services more sporadically.

Although we reported a high linkage rate (92.3%), we previously described a linkage bias showing certain characteristics such as age, gender, and ethnicity were associated with an increased risk of not being successfully linked 11]. Another limitation is that our sample only contains patients who have been referred to SLaM, meaning we cannot compare our findings to a population with mental health disorders who have not accessed secondary mental health services. Further, we do not hold reliable data across the cohort specifically indicating whether a patient is currently in or out of work, though there are indicators among those receiving UC as to whether someone is currently in, out of, or looking for work.

This data linkage provides an opportunity to gain extensive knowledge around complex relationships between benefit receipt and mental health as well as provide evidence to support and answer areas of research interest stated by the DWP relating to health and disability and welfare provision 2
15], and around mental health provision. Future work will initially look at temporality of benefit receipt and mental health diagnosis, as well as the amount and type of interaction patients have with mental health services and the benefits system.

### Data access

The linked data is not freely available due to ethical and legal restrictions that exist for the current data linkage to ensure the privacy of patients. The team welcomes research enquiries for project proposals and collaboration. Those who are interested should contact the study lead, Dr Sharon Stevelink who will advise on the practicality, processes and required permissions for using the data linkage.

### Author contributions

AP, SAMS and NTF conceptualised the design of this cohort profile study. MH, JD, NTF, SD and SAMS were part of initial discussions, the various processes and then finalization of the data linkage itself. MB, RL and AJ took lead in data curation. AP led on project administration and analysis, wrote initial draft of the paper including all sections and revised the paper. SAMS, NTF, MH and SD provided supervision and supported on drafts of the paper. RL provided data cleaning and statistical support. AP and SAMS acquired funding for the study with support from NTF and MH. All authors commented on final draft of the paper. NTF and SAMS are joint last authors.

### Patient and public involvement statement

This project was informed by discussions with the NIHR Maudsley BRC Data Linkage Service User and Carer Advisory Group.

### Patient consent for publication

Not required.

## Data Availability

The linked data is not freely available due to ethical and legal restrictions that exist for the current data linkage to ensure the privacy of patients (see ethics statement below). The team welcomes research inquiries for project proposals and collaboration. Those who are interested should contact the study lead, Dr Sharon Stevelink who will advise on the practicality, processes and required permissions for using the data linkage. Data may be obtained from a third party and are not publicly available. Data are not publicly available. Access to deidentified data can be applied for via the NIHR Maudsley Biomedical Research Centre at the South London and Maudsley NHS Foundation Trust, upon reasonable request. Requests for data will be considered on a case-by-case basis, given the sensitive nature of the data, and access will only be granted if approval is given by the Work and Health Screening Panel and other governance requirements are fulfilled. For more information, please contact. Ethics statement:The South London and Maudsley (SLaM) NHS Foundation Trust and Department for Work and Pensions (DWP) data linkage resulted in a linked data set of longitudinal data from SLaM electronic mental healthcare records and DWP benefits data. The data linkage was completed in late 2020 and thereafter available for research purposes. All data used is de-identified. The use of SLaM mental healthcare records data for research purposes received full ethical approval from the NHS Research Ethics Committee (Oxford South Central ref 18/SC/0372). Ethical approval for the data linkage was granted from the Oxford South Central REC (Ref: 22/SC/0400), and the data linkage was legally permitted by the Health Research Authority Confidentiality Group (CAG) under Section 251 of the NHS Act 2006 (ref 17/CAG/0055).

## Acknowledgements

We would like to thank Megan Pritchard at the National Institute for Health and Care Research (NIHR) Maudsley Biomedical Research Centre (BRC) for their support with this study. We would also like to thank the members of the NIHR Maudsley BRC Data Linkage Service User and Carer Advisory Group for their input. We are very grateful to the DWP/Department of Health and Social Care joint Work and Health Directorate staff, who supported us in creating this linked dataset and for all the advice provided.

## References

1. Department for Work and Pensions. Work, Health and Disability Green Paper Data Pack. 2016. https://assets.publishing.service.gov.uk/government/uploads/system/uploads/attachment_data/file/644090/work-health-and-disability-green-paper-data-pack.pdf (March 2023, date last accessed).

2. Department for Work and Pensions. Sharping future support: the health and disability green paper. August 2021. https://www.gov.uk/government/consultations/shaping-future-support-the-health-and-disability-green-paper (March 2023, date last accessed).

3. Department for Work and Pensions. Benefits and financial support if you’re temporarily unable to work. https://www.gov.uk/browse/benefits/unable-to-work (March 2023, date last accessed).

4. Department for Work and Pensions. Universal Credit. https://www.gov.uk/universal-credit/what-youll-get. (March 2023, date last accessed).

5. Department for Work and Pensions. From Welfare state to welfare system. November 2016. https://www.gov.uk/government/speeches/from-welfare-state-to-welfare-system (March 2023, date last accessed).

6. Dwyer P, Scullion L, Jones K, McNeill J, Stewart AB. Work, welfare, and wellbeing: The impacts of welfare conditionality on people with mental health impairments in the UK. Social Policy & Administration. 2020 Mar;54(2):311–26.

7. Wickham S, Bentley L, Rose T, Whitehead M, Taylor-Robinson D, Barr B. Effects on mental health of a UK welfare reform, Universal Credit: a longitudinal controlled study. The Lancet Public Health. 2020 Mar 1;5(3):e157–64.

8. Jewell A, Pritchard M, Barrett K, Green P, Markham S, McKenzie S, Oliver R, Wan M, Downs J, Stewart R. The Maudsley Biomedical Research Centre (BRC) data linkage service user and carer advisory group: creating and sustaining a successful patient and public involvement group to guide research in a complex area. Research involvement and engagement. 2019 Dec;5(1):1–0.

9. NHS South London and Maudsley NHS Foundation Trust. Who we are. https://slam.nhs.uk/about (March 2023, date last accessed).

10. Stewart R, Soremekun M, Perera G, Broadbent M, Callard F, Denis M, Hotopf M, Thornicroft G, Lovestone S. The South London and Maudsley NHS foundation trust biomedical research centre (SLAM BRC) case register: development and descriptive data. BMC psychiatry. 2009 Dec;9(1):1–2.

11. Stevelink SA, Phillips A, Broadbent M, Boyd A, Dorrington S, Jewell A, Leal R, Bakolis I, Madan I, Hotopf M, Fear NT. Linking electronic mental health and benefits records in South London: design, procedure and descriptive outcomes. BMJ Open. 2023 Feb 1;13(2):e067136.

12. Department for Work and Pensions. State Pension age timetable. May 2014. https://www.gov.uk/government/publications/state-pension-age-timetable/state-pension-age-timetable (April 2023, date last accessed).

13. Kennedy S. The Work Capablity Assessment for Employment and Support Allowance. November 2012. https://researchbriefings.files.parliament.uk/documents/SN05850/SN05850.pdf (March 2023, date last accessed).

14. McManus S, Bebbington PE, Jenkins R, Brugha T. Mental health and wellbeing in England: the adult psychiatric morbidity survey 2014. NHS Digital; 2016.

15. Department for Work and Pensions. DWP Areas of Research Interest. March 2019. https://www.gov.uk/government/publications/dwp-areas-of-research-interest-2019 (March 2023, date last accessed).

